# Comparison of Cerebral ECD Perfusion in Patients with Dementia with Lewy Bodies and Parkinson’s Disease Dementia

**DOI:** 10.1101/2024.10.02.24314782

**Authors:** Aili Toyli, Guang-Uei Hung, Chen Zhao, Qiuying Sha, Pai-Yi Chiu, Weihua Zhou

**Affiliations:** Department of Mathematical Sciences, Michigan Technological University, Houghton, MI, USA; Department of Nuclear Medicine, Chang Bing Show Chwan Memorial Hospital, Changhua, Taiwan; Department of Computer Science, Kennesaw State University, Marietta, GA, USA; Department of Neurology, Show Chwan Memorial Hospital, Changhua, Taiwan; Department of Applied Computing, Michigan Technological University, Houghton, MI, USA; Center for Biocomputing and Digital Health, Institute of Computing and Cyber-systems, and Health Research Institute, Michigan Technological University, Houghton, MI, USA

## Abstract

Dementia with Lewy bodies (DLB) and Parkinson’s Disease Dementia (PDD) are closely related neurodegenerative conditions within the Lewy body spectrum. The relationship between DLB and PDD remains debated, with ongoing discussion about whether they are distinct diseases or different manifestations of the same disorder. This study aimed to identify differences in cerebral perfusion patterns between DLB and PDD patients. Single-photon emission computed tomography (SPECT) Ethyl Cysteinate Dimer (ECD) imaging was performed on each patient, and relative tracer uptake levels across 47 regions of interest (ROIs) and 240 subregions were analyzed. A two-sided Welch’s t-test was employed to evaluate mean perfusion differences, with results further confirmed through a voxel-wise t-test mapping. After adjusting for multiple comparisons using the Benjamini-Hochberg procedure, no regions showed statistically significant differences at the 0.05 level. However, a few subregions in the visual cortices exhibited p-values just above the significance threshold, with lower mean perfusion observed in PDD patients than those with DLB. Validating these findings in larger samples could enhance scientific understanding of the differences in the pathology and progression of DLB and PDD.

## Background

Dementia with Lewy Bodies (DLB) and Parkinson’s Disease Dementia (PDD) are two forms of Lewy body syndrome presenting similar clinical and neuropathological features [1]. The scientific community has long considered the possibility that they are two presentations of the same underlying disease. The current standard for differentiation is known as the “one-year rule” [2]. DLB is diagnosed if cognitive impairment emerges before or within one year of parkinsonian motor signs, whereas PDD is diagnosed if Parkinson’s Disease (PD) symptoms are established before cognitive impairment. Both DLB and PDD progress with similar patterns of impairment and rates of cognitive decline [3]. Postmortem analyses reveal similar distributions and severities of Lewy body pathology in DLB and PDD [1]. A notable difference between the two diseases is the higher incidence of amyloid plaques and Alzheimer’s disease (AD) co-pathology in DLB [4], which some argue is responsible for the earlier onset of dementia relative to parkinsonism [1].

Given the clinical and neuropathological similarities between DLB and PDD, much work has been done to distinguish them using neuroimaging techniques. Comparisons of brain magnetic resonance imaging (MRI) scans show similar patterns of atrophy, with the only notable distinction being more significant impairment in the pallidus for DLB patients [5, 6]. A multitracer positron emission tomography (PET) study did not find significant differences between DLB and PDD [7]. Other studies have used single-photon emission computed tomography (SPECT) to visualize and compare brain perfusion for DLB and PDD patients [6, 8-10]. Many of these studies utilized the technetium-99m-ethyl cysteinate dimer (ECD) tracer, preferred for its high brain-to-soft-tissue activity ratio and helpful for identifying regions with hypoperfusion [11, 12]. Three previous studies [8-10] found no statistically significant difference in perfusion. Another study identified higher cerebral blood flow (CBF) in the whole cingulate gyrus and lower CBF in the precuneus area in DLB than PDD, but no level of statistical significance was provided [6].

Previous studies compared ECD SPECT scans for PDD and DLB using either manually determined regions of interest (ROIs) or voxel-wise comparison [6, 8-10]. In this study, we performed an analysis using regional tracer uptake values of ECD for computer-generated ROIs. While voxel-wise comparison identifies perfusion differences in highly specific regions, ROI analysis compares labeled anatomical regions, offering greater interpretability for clinicians. Automatically determined ROIs also provide higher reliability and reproducibility than those determined manually [13]. We aimed to determine whether comparing the average tracer uptake values calculated would identify statistically significant differences and corroborate regions previously identified to have perfusion differences between PDD and DLB patients.

## Cohort

The cohort consisted of 30 patients with diagnosed PDD and 46 with DLB. Patients with DLB were diagnosed according to the revised consensus criteria for probable DLB, developed by the fourth report of the DLB consortium [14]. The PDD patients were diagnosed according to the clinical criteria for PD developed by the Movement Disorder Society (MDS) in 2015 [15] and for probable PDD by the MDS in 2007 [16].

Demographic information for DLB and PDD cohorts can be viewed in Table 1. The DLB cohort was slightly older. The sex ratio was approximately equal for both groups. The results of several clinical assessments of dementia are also shown. Note that both cohorts scored similarly, with the PDD group having a lower CDR-SB and a higher MoCA score. The PDD cohort had a higher LEDD, which is reasonable as levodopa is the basis of pharmacological treatment for PD.

**Table 1.** Demographic information for DLB and PDD cohorts is shown. Summaries of sex breakdown and patients’ ages is accompanied by scores on numerous cognitive assessments. P-values are shown of a Fisher chi-squared test for sex and t-tests for equality of the means without equal variance assumed for all other variables. The number of patients in each cohort is listed for variables for which data was not collected for the entire cohort. (Abbreviations: CDR-SB: sum of boxes of the Clinical Dementia Rating [17], MoCA: Montreal Cognitive Assessment [18], NPI: Neuropsychiatric Inventory [19], HAIADL: Activities of Daily Living scale in the History-based Artificial Intelligent Clinical Dementia Diagnostic System [20], UPDRS III: part 3 of the Unified Parkinson’s Disease Rating Scale [21], LEDD: levodopa equivalent dose [22], SBR: striatal background ratio).

|  | DLB | PDD | p-value |
| --- | --- | --- | --- |
| Number of patients | 46 | 30 |  |
| Age (Years) | 78 +/- 2.3 | 74 +/- 3.6 | 0.077 |
| Education | 3.7 +/- 1.3 | 6.7 +/- 1.6 | 0.006 |
| Sex | 50% Male,<br>50% Female | 53.3% Male,<br>46.7% Female | 0.776 |
| CDR (# of patients) |  |  |  |
| 0.5 | 9 | 13 |  |
| 1 | 20 | 11 |  |
| 2 | 16 | 4 |  |
| 3 | 1 | 2 |  |
| CDR-SB | 7.9 +/- 1.2 | 6.6 +/- 1.7 | 0.217 |
| MoCA (45 DLB, 28 PDD) | 7.7 +/- 1.8 | 10.8 +/- 2.9 | 0.079 |
| NPI | 12.7 +/- 4.4 | 9 +/- 4.5 | 0.248 |
| HAIADL | 14.6 +/- 2.1 | 11.9 +/- 3.0 | 0.145 |
| UPDRS III (23 DLB, 8 PDD) | 33.5 +/- 3.7 | 34.7 +/- 13.1 | 0.873 |
| LEDD (43 DLB, 28 PDD) | 218.2 +/- 67.6 | 383.7 +/-<br>115.1 | 0.019 |
| SBR (16 DLB, 19 PDD) | 1.36 +/- 0.20 | 1.19 +/- 0.31 |  |

## Methods

The data were acquired by two SPECT/CT systems (Infinia Hawkeye, GE Healthcare, Wauwatosa, WI, USA; Symbia, Siemens Healthineers, Erlangen, Germany) from 2 affiliated centers (Show Chwan Memorial Hospital; Chang Bing Show Chwan Memorial Hospital) and were retrospectively recruited under the local ethics approval. See detailed imaging acquisition parameters in Table 2.

**Table 2.** Imaging acquisition parameters of systems utilized for SPECT ECD scans at each data collection center.

|  |  | Chang Bing Show Chwan | Show Chwan |
| --- | --- | --- | --- |
| Camera |  | GE-Infinia-Hawkeye | SIEMENS Symbia TruePoint SPECT/CT |
| Workstation |  | Xeleris | Esoft ICS |
| Time to imaging (after injection) |  | 30 mins | 30 mins |
| Acquisition | Injection dose | 30mCi | 30mCi |
|  | Collimator | LEHR | LEHR |
| | Energy windows | 140 keV±10% | 140 keV±15% |
|  | Sampling angle | 6 degree | 5.6 degree |
|  | Acquisition type | Step | Step and shoot |
|  | Acquisition time | 25s/view | 30s/view |
|  | Matrix size | 128*128 | 128*128 |
|  | Pixel size | 0.83mm | 2.7mm |
|  | Acquisition zoon | 1.33 | 1.78 |
| Reconstruction | Pre-filtering | OSEM/MLEM<br>Number of OSEM iterations: 5<br>Max number of subsets: 6 | - |
|  | Post-filtering | Butterworth<br>Critical Frequency: 0.5<br>Power: 10 | Flash: 3D<br>Subsets: 4<br>Iterations: 6 |
|  | Scatter correction | on | on |

All scans were uploaded into NeuroQ (NeuroQ v3.80; Syntermed, Inc.; Atlanta, GA) and normalized to the software’s normal ECD template using twenty iterations. The whole brain was selected as the reference region to normalize uptake values to the average pixel value among all regions in each scan. Relative uptake values for 47 ROIs and 240 subregions divided by horizontal plane for each scan were calculated.

A two-sided Welch’s t-test on mean relative perfusion for each ROI for DLB and PDD patients was performed in R (R Core Team, 2023; R Foundation for Statistical Computing; Vienna, Austria.). The Benjamini-Hochberg procedure was used to correct the p-value for the inflated false discovery rate associated with multiple tests [23]. P-value adjustment was performed on results from the 47 ROIs and separately for the 240 subregions.

After comparing relative perfusion values derived from NeuroQ, test results were verified and visualized using the open-source SPM12 software for MATLAB (SPM12 v7771; Wellcome Trust Centre for Neuroimaging, Institute of Neurology; University College London, UK). They were normalized to the built-in SPM SPECT template and smoothed using default settings. A two-sided voxel-wise t-test map was generated in SPM and the results were rendered for 3-dimensional visualization.

## Results

No ROI showed a significant difference in relative perfusion after p-value adjustment. DLB and PDD patients followed similar patterns of hypoperfusion. A few subregions had p-values only slightly greater than the pre-established alpha = 0.05 significance level (see Table 3). Adjusted p-values were over 0.1 for all other ROIs.

**Table 3.**
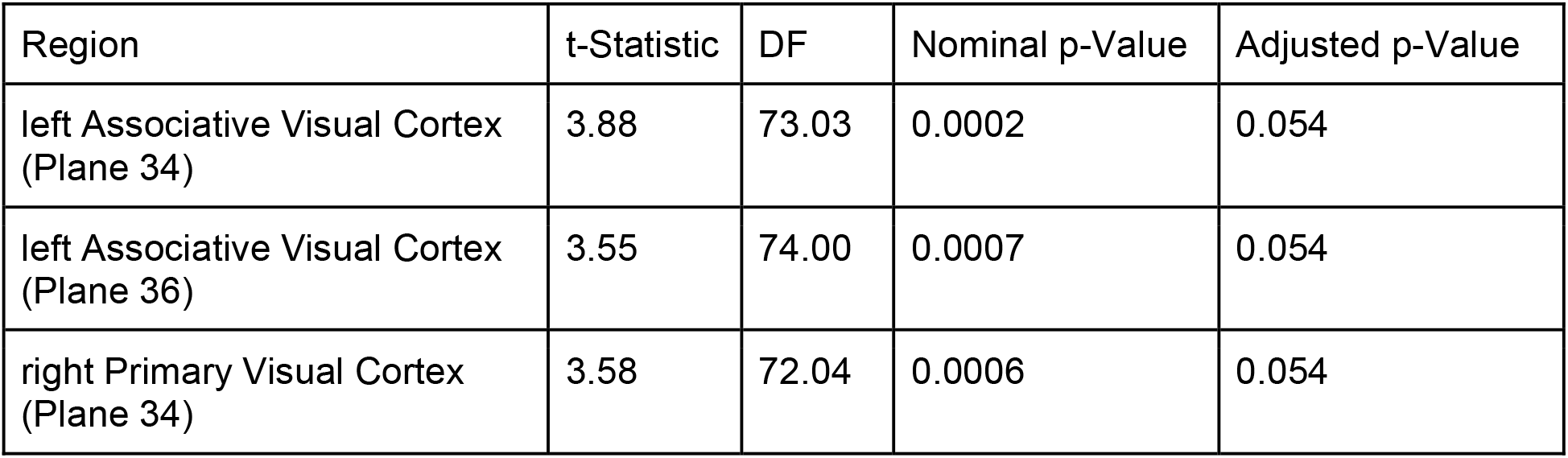
Results of two-sides Welch’s t-test on relative ECD perfusion difference between DLB and PDD cohorts. The regions with the lowest p-values are shown. Note that these ROIs are all located in the visual cortices and have adjusted p-values just above the significance level of 0.05. All other ROIs had adjusted p-values over 0.1.

The regions in which the two groups had the greatest difference in perfusion were in the visual cortices, where perfusion was lower for patients with PDD than those with DLB. Side-by-side comparison of two example patients chosen to highlight this difference is demonstrated in Figure 1.

**Figure 1.**
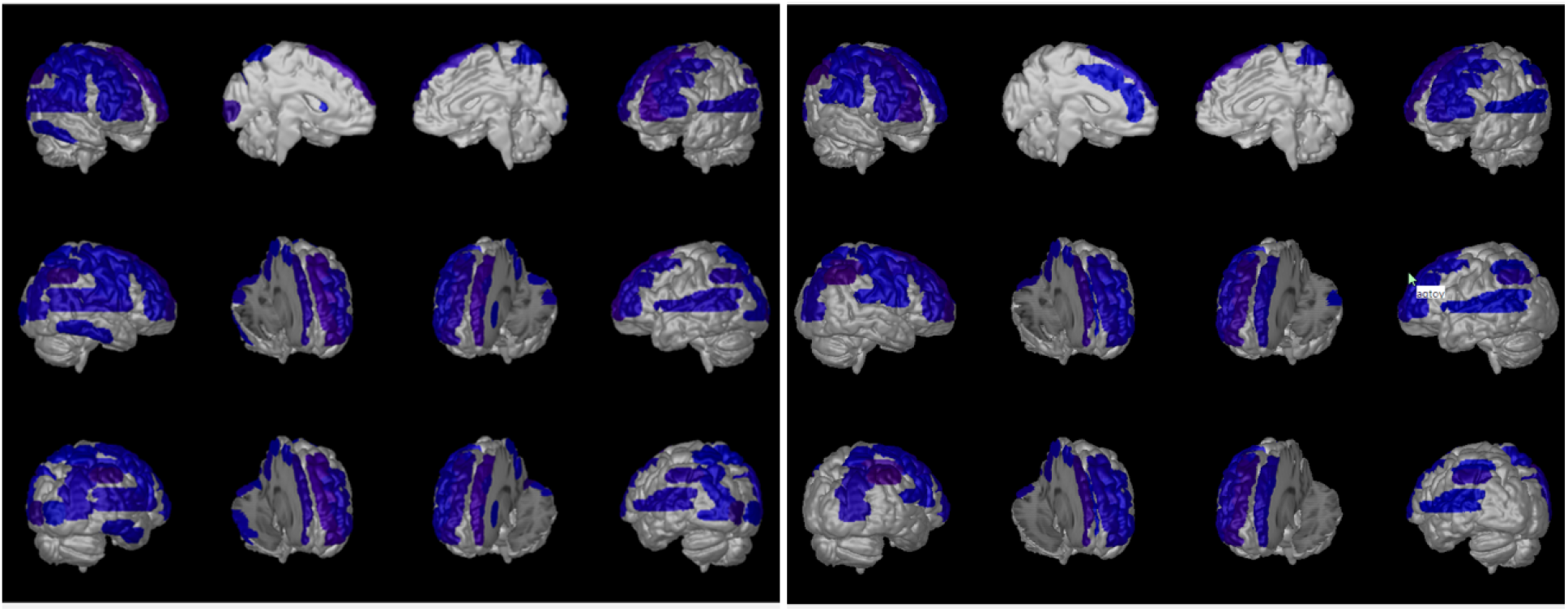
Side-by-side comparison of PDD (left) and DLB (right) example patients. Visuals generated with NeuroQ software. Blue indicates regions of relative hypoperfusion. Overall patterns of hypoperfusion are similar for both patients, with additional hypoperfusion in the occipital lobe for the PDD patient.

**Figure 2.**
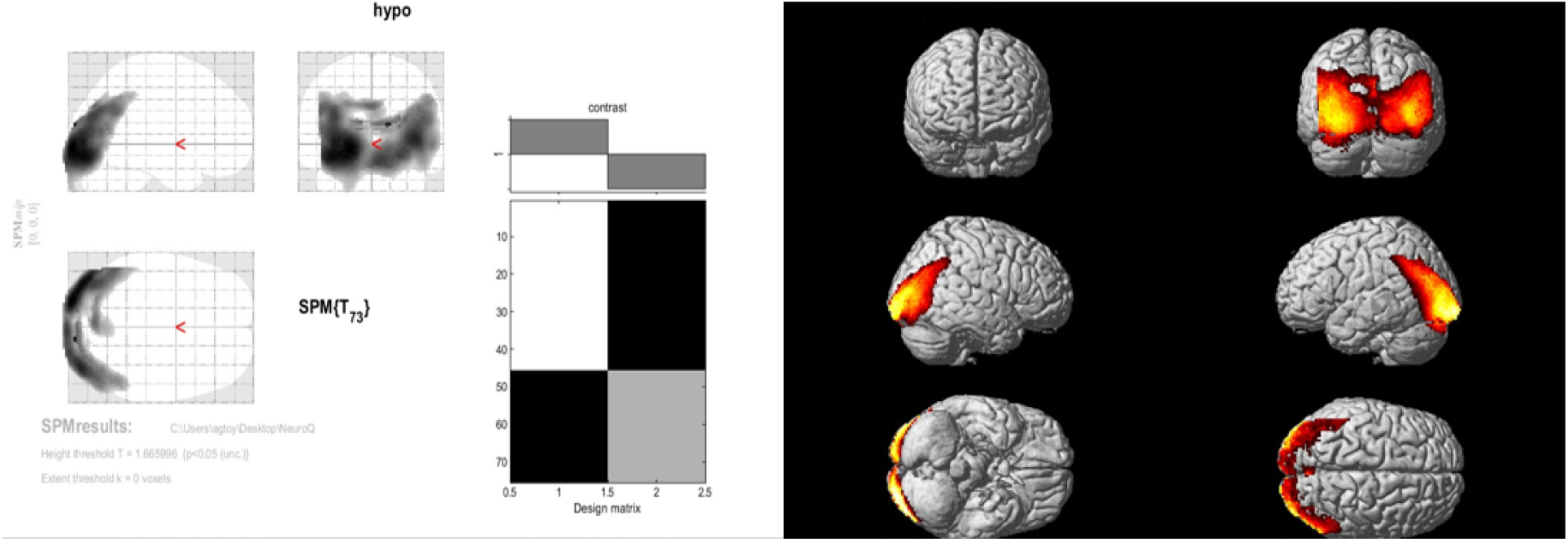
SPM12 generated t -map comparing relative perfusion of mean images of DLB and PDD scans with a nominal significance level of 0.05. Highlighted areas in the occipital lobe indicate greater relative hypoperfusion in PDD. No regions were found with greater hypoperfusion in DLB. Note: No differences remained significant after p-value adjustment.

Voxel-wise results corroborated ROI differences. The visual cortices showed the greatest reduction in relative perfusion in PDD compared to DLB, and no regions were found with a greater reduction in DLB.

## Discussion

Although no ROI or subregion comparison of DLB and PDD passed the predetermined statistical significance threshold, it is notable that comparisons of the left-associative visual cortex on planes 34 and 36 and the right primary visual cortex on plane 34 had p-values just above the adjusted significance level. It may still be worthwhile to explore these differences further, especially considering our small sample size. We followed the standard practice and used a significance level of 0.05. However, marginally insignificant p-values, such as those less than 0.1, may indicate a noteworthy relationship [24]. Future studies with a larger cohort would provide higher power of the test and may provide statistically significant results where we and previous studies could not [8,9].

Some previous studies have indicated greater reduced perfusion in the visual cortices or in the occipital lobe for PDD patients. Takemoto et al. created a subtracted image of mean intensity values of PDD (n = 52) and DLB (n = 46) patients [6]. Their subtracted image shows reduced CBF in the left visual cortex. Rossi et al. used SPM2 to detect significant perfusion reduction in the occipital lobes of DLB (n = 30) and PDD (n = 30) patients compared to healthy controls or AD patients. Still, there were no significant differences between the DLB and PDD cohorts [8]. Another study by Mito et al. using 3D-SSP for pixel-wise comparison found cerebral perfusion was slightly decreased at the lateral parietal association, lateral temporal association, posterior cingulate cortex, and precuneus in DLB (n = 6) patients compared to those with PDD (n = 7) [9]. The difference in results between the aforementioned study and the present one could be explained by variation induced by the small sample size.

The hypoperfusion pattern identified may be associated with the clinical progression of DLB and PDD. Occipital hypoperfusion in DLB and PDD are both associated with better cognitive function [25, 26]. In this study, PDD participants had better cognitive functions than DLB according to performance on MoCA and less severe dementia stage according to the CDR-SB. We think this was the main reason why there was more reduced perfusion in the occipital lobe for PDD. In addition, we hypothesize that subcortical regions including striatal, thalamic, and cerebellar areas share more blood supply than occipital lobe when motor dysfunction is getting worse. This concept is based on compensatory perfusion to more dysfunctional regions via the same blood supply from posterior circulation of the brain. PDD usually had more motor dysfunction than DLB. We think this may also explain why PDD had more occipital hypoperfusion.

Several limitations of this study must be addressed. First is the failure to account for covariates affecting disease progression and relative perfusion. Factors such as age, time from onset of disease, and AD co-pathology have been shown to affect the progression of PDD and DLB [1]. AD co-pathology is considered one of the most notable differences between DLB and PDD, more frequently occurring in DLB [27]. Some consider AD co-pathology to be the underlying difference between DLB and PDD [1]. However, not all DLB patients have an AD diagnosis [27].

Another important limitation is our assessment of relative perfusion. All tracer uptake values were standardized by the whole-brain average uptake value to account for the difference in tracer uptake between individuals. All uptake values are relative measures and cannot be used as an absolute comparison of perfusion between individuals.

Additionally, the t-test identifies differences in mean relative perfusion. The potential differences identified between groups cannot necessarily be used to differentiate between individual scans or diagnoses. However, regional differences in relative perfusion can be used to better understand the difference between the pathology of these two diseases, aiding future research into their causes and progression.

## Conclusion

No statistically significant differences in ECD perfusion were found between DLB and PDD patients using Welch’s t-test. The greatest difference was in the visual cortices, where PDD patients suffered a greater loss of perfusion. Future research with a larger cohort may significantly confirm these findings.

## Data Availability

All data produced in the present study are available upon reasonable request to the authors

